# Exome-wide tandem repeats confer large effects on subcortical volumes in UK Biobank participants

**DOI:** 10.1101/2023.12.11.23299818

**Authors:** Mary Anne Panoyan, Yuxin Shi, Cristina L Abbatangelo, Nina Adler, Ashley Moo-Choy, Esteban J Parra, Renato Polimanti, Pingzhao Hu, Frank R Wendt

## Abstract

The human subcortex is involved in memory and cognition. Structural and functional changes in subcortical regions is implicated in psychiatric conditions. We performed an association study of subcortical volumes using 15,941 tandem repeats (TRs) derived from whole exome sequencing (WES) data in 16,527 unrelated European ancestry participants. We identified 17 loci, most of which were associated with accumbens volume, and nine of which had fine-mapping probability supporting their causal effect on subcortical volume independent of surrounding variation. The most significant association involved *NTN1*-[GCGG]_N_ and increased accumbens volume (β=5.93, P=8.16x10^-9^). Three exonic TRs had large effects on thalamus volume (*LAT2*-[CATC]_N_ β=-949, P=3.84x10^-6^ and *SLC39A4*-[CAG]_N_ β=-1599, P=2.42x10^-8^) and pallidum volume (*MCM2*-[AGG]_N_ β=-404.9, P=147x10^-7^). These genetic effects were consistent measurements of per-repeat expansion/contraction effects on organism fitness. With 3-dimensional modeling, we reinforced these effects to show that the expanded and contracted *LAT2*-[CATC]_N_ repeat causes a frameshift mutation that prevents appropriate protein folding. These TRs also exhibited independent effects on several psychiatric symptoms, including *LAT2*-[CATC]_N_ and the tiredness/low energy symptom of depression (β=0.340, P=0.003). These findings link genetic variation to tractable biology in the brain and relevant psychiatric symptoms. We also chart one pathway for TR prioritization in future complex trait genetic studies.

## INTRODUCTION

The subcortical brain comprises the accumbens, amygdala, caudate, hippocampus, pallidum, putamen, and thalamus. These regions play significant roles in cognitive and emotional functions.^1^ Variation in subcortical volume may affect pathology of cognitive, psychiatric, and movement disorders.^1^ For example, schizophrenia is associated with decreased volume in the hippocampus, amygdala, and thalamus, but increased volume in the bilateral caudate, putamen, and pallidum,^2^ depression is associated with reduced hippocampus volume,^3^ panic disorder patients have reduced thalamus volume relative to controls,^4^ and posttraumatic stress disorder (PTSD) is associated with reduced volume of the amygdala.^5^ In addition to their association with mental health symptoms and diagnoses, subcortical volumes may vary due to factors such as sex, aging, lifestyle, and genetics.^6–8^ Recent studies have identified single nucleotide polymorphisms (SNPs) from genome-wide association studies (GWAS) influencing subcortical volumes ^8–10^. Twin heritability estimates for brain volumes range from 34-95%.^8,11^ SNP-based heritability estimates in a European sample ranged from 17% for the amygdala to 47% for the thalamus using genome-wide complex trait analysis, and ranging from 9% for the amygdala to 33% for the brainstem using linkage disequilibrium score regression (LDSC).^8^ Despite a prominent genetic component to subcortical volume variation and a consistent relationship between subcortical structural and functional abnormalities and various psychiatric outcomes, we still have limited understanding of the pleiotropic mechanisms linking them. Some notable consistencies between genetics and phenotypic studies include a genetic correlation (r_g_) between schizophrenia and reduced hippocampus volume (r_g_=-0.18)^12^ and between anxiety with reduced putamen volume (r_g_=-0.482).^13^

Current GWAS only target SNPs and short insertion/deletion polymorphisms to characterize the genetics of subcortical volumes. Another class of genetic variation, tandem repeats (TRs), are genomic loci that consist of consecutively repeating motifs, typically 1-20 base pairs in length. They have relatively high mutation rates (10^-5^ to 10-^2^) compared SNPs (10^-8^), are multiallelic, and when localized to regulatory or coding regions, can inform actionable biology associated with subcortical brain volume.^14^ Recent studies have demonstrated the utility of TR-based studies to uncover novel complex-trait associated loci that are independent of surrounding SNPs discovered in GWAS.^15–17^ Analysis of TR variation associated with subcortical volumes presents a unique opportunity for uncovering novel insights into the genetic basis of brain structure and function.

We conducted exome-wide association studies for seven subcortical brain volumes using 15,941 TRs and 21,538 individuals with European ancestry from the UKB. After applying false discovery rate to account for multiple testing correction (FDR<5%), we identified 17 TRs associated with subcortical volumes, highlighting the role of specific genes in brain structural changes. A subset of these TRs appear to have large effects on brain biology that can lead to symptoms of major depression, generalized anxiety, and posttraumatic stress. These findings reveal potentially actionable variation in the genome that links brain volume to psychiatric symptoms in a way that traditional GWAS have not. In doing so, we chart one path forward for uncovering novel and biologically relevant genetic signals underlying mental health and wellbeing.

## METHODS

### UK Biobank Subcortical Volumes

The UKB is a large population-based study of over 500,000 participants. Trait assessment in the UKB includes a range of factors such as diet, behavior, physical activity, mental health, etc.. A subset of individuals underwent brain imaging using magnetic resonance imaging (MRI) to estimate volume of the accumbens, amygdala, caudate, hippocampus, pallidum, putamen, and thalamus. T1-weighted structural imaging was used to depict high-resolution images of these brain regions. Images were taken with a Siemens Skyra 3T running VD13A SPA with standard 32-channel RF received hear coil. T1-weighted structural imaging was performed over a five-minute timeframe with 1x1x1 mm resolution and a 208x256x256 field-of-view matrix. Subcortical volumes were modeled from T1 data using FIRST (FMRIB’s Integrated Registration and Segmentation Tool).^18^ Fourteen subcortical volumes were generated from T1 images; two volumes per region representing left and right volumetric measurements in cubic millimeters (mm^3^). In this study, we summed left and right hemisphere volumetric measurements to assess a single volume per subcortical region.

UKB participants were assigned to one of six ancestry groups using genetic principal components and a random forest classifier based on the 1000 Genomes Project Phase 3 and Human Genome Diversity Panel ancestry groups.^19^ See the Pan-UKB webpage for additional details: https://pan.ukbb.broadinstitute.org on this procedure. Because of the limited sample size available in other population groups, our discovery cohort used 16,527 unrelated European ancestry participants from the UKB. As mentioned below, information regarding other ancestry groups (182 African, 53 admixed American, 349 Central/South Asian, 179 East Asian, and 58 Middle Eastern participants) was used in the replication analyses. This research has been conducted in the scope of UKB application reference number 58146.

### UK Biobank Tandem Repeat Association Testing

UKB CRAM files of WES reads aligned to the hg38 reference genome were converted to binary alignment map (BAM) files using cramtools v3.0 (February 2021).^20^ Genotyping of autosomal tandem repeats from short reads was performed with GangSTR v2.5.0^21^ using sorted and indexed BAMs. To associate each multi-allelic TR with subcortical volumes, TR genotypes were converted to a locus-level burden of allele lengths by summing both TR alleles in a genotype. In R, generalized linear models were used to regress TR burden on each subcortical volume using age, sex, sex×age, age^2^, sex×age^2^, and the first 10 within-ancestry principal components as covariates. The test statistics from TR association models of each trait were evaluated for systematic biases by estimating genomic inflation factor (lambdaGC) with the following lines of code executed in R Studio:

`chisq <- qchisq(1-data$pval,1)`

`lambdagc <- median(chisq)/qchisq(0.5,1)`

Multiple testing correction was applied two ways. First, using an FDR of 5% per subcortical region to assess trait-level significance. Second, using a genome-wide multiple testing correction commonly used in SNP-based genome-wide association studies (P<5x10^-8^). We replicated significantly associated loci using five additional cohorts all containing unrelated participants: 182 African, 53 admixed American, 349 Central/South Asian, 179 East Asian, and 58 Middle Eastern participants.

### Locus Annotation Overview

TRs were functionally annotated for the following features/effects: (i) fine-mapping of TR-containing genomic regions relative to surrounding bi-allelic variation, (ii) phenome-wide association study (PheWAS) to identify pleiotropic effects, (iii) gene set enrichment of discovered loci, (iv) gene expression and splicing effects, (v) population genetics framework to infer selection at short TRs (SISTR),^22^ and (vi) model protein folding effects of exonic TRs using AlphaFold.^23^ These procedures are described below.

### Phenome-wide association of TR-containing genes

TRs were positionally mapped to genes using the human hg38 reference genome. Hypergeometric tests were used to determine the enrichment of pleiotropic associations of each gene using results from the GWAS Atlas PheWAS feature (https://atlas.ctglab.nl/PheWAS) which included 4,756 traits separated into 27 trait domains: activities (N=137), aging (N=5), body structures (N=18), cardiovascular (N=162), cellular (N=1,143), cognitive (N=78), connective tissue (N=14), dermatological (N=31), ear, nose, and throat (N=6), endocrine (N=67), environmental (N=92), gastrointestinal (N=42), hematological (N=38), immunological (N=365), infection (N=3), metabolic (N=1,259), mortality (N=46), muscular (N=5), neoplasms (N=49), neurological (N=437), nutritional (N=46), ophthalmological (N=28), psychiatric (N=321), reproduction (N=76), respiratory (N=74), skeletal (N=194), and social interactions (N=20).

### Gene set enrichment

Pathway enrichment was performed using three different methods; g:Profiler,^24^ ShinyGO v0.77,^25^ and Enrichr.^26^ Enrichment with g:Profiler was performed with an ordered query as input (i.e., genes were ranked in order based on p-value for the contained TR) with otherwise default parameters being used. ShinyGO and Enrichr enrichments were performed using default parameters. The significant pathways were visualized using Cytoscape^27^ with the colour of the nodes reflecting the method from which the pathways reached significance. Tissue enrichment with Multi-marker Analysis of GenoMic Annotation (MAGMA)^28^ was implemented in Functional Mapping and Annotation of GWAS (FUMA)^29^ using 54 specific Genotype-Tissue Expression (GTEx) v8 tissues, 11 general developmental stages of brain samples from BrainSpan, and 29 specific ages of brain samples from BrainSpan.

### Gene expression and splicing effects of TRs

Gene expression-associated TRs (eSTR) and splicing-associated TRs (spl-TRs) were identified using data from Fotsing, et al.^30^ and Hamanaka, et al.,^31^ respectively. Briefly, both reports have made publicly available the associating statistics between TR variation and gene expression or splicing. Both resources used the Genotype-Tissue Expression Project including all v7 individuals for eSTRs (17 tissues studied in N=652 unrelated donors; 86% European ancestry) and v8 for spl-TRs (49 tissues studied in N=838 unrelated donors; 84.6% European ancestry). For eSTRs, average TR repeat lengths were called from hg19-aligned whole genome sequences using HipSTR.^32^ For spl-TRs, average TR repeat lengths were called from hg38-aligned whole genome sequences using GangSTR.^21^ Both studies used TR dosage as a predictor of expression or splicing quantity. The eSTR models included age, sex, and the first 10 within-ancestry principal components as covariates. The spl-TR models included sex, library preparation protocol, sequencing platform, the first five within-ancestry principal components, and probabilistic estimation of expression residuals.

### Selection inference at TR loci

SISTR^22^ measures negative selection against individual TR alleles by fitting an evolutionary model of TR variation that includes mutation, genetic drift, and negative natural selection to empirical allele frequency data to infer the posterior distribution of selection coefficients (*s*) at TRs. The *s* parameter reflects a decrease in reproductive fitness for each repeat unit away from the most frequent allele (i.e., the modal allele) at a locus. SISTR supports *s* inference for TRs with 2-4 basepair motifs due to their abundance in the genome and reliable mutation models. Other repeat motif lengths (e.g., homopolymers and those with ≥5 basepairs) are less abundant, may follow mutation models different from the generalized stepwise model used by SISTR, may have low mutation rates, and/or have noisy population-level allele frequency estimates. SISTR assumes that the most frequent allele in a present-day population has optimal evolutionary fitness, the fitness landscape per locus is symmetric around the most frequent allele, and fitness effects are additive. Allele frequencies per TR were estimated using the same European ancestry samples used to perform TR-trait association.

### Fine-mapping of TR-containing genomic regions

We utilized the program FINEMAP (version 1.4.2)^33^ to detect candidate causal variants from all associated regions. FINEMAP employs a Bayesian framework that leverages summary association data and linkage disequilibrium (LD) among variants to calculate the posterior probability of causality using a shotgun stochastic search algorithm.^33^ The regions selected for fine-mapping were defined as ± 500 kb on either side of the lead TR. LD matrices and SNP effect sizes were calculated using the same set of UKB individuals used for TR association testing. For each locus, the maximum number of causal variants was set to 10 (i.e., a maximum of 10 credible sets). A credible set is comprised of variants that cumulatively reach a probability of at least 95%. The variants within a credible set are referred to as candidate causal variants and each of them has a corresponding posterior inclusion probability (PIP). FINEMAP results were filtered by removing candidate causal variants with a log_10_(Bayes Factor (BF)) > 2 and p-value < 10^-5^ from each of the 95% credible sets. The Bayes Factor compares the likelihood of two competing hypotheses: 1) the variant is causally associated with the phenotype, and 2) the variant is not associated with the phenotype. Thus, the log_10_BFs represent the strength of evidence supporting one hypothesis over the other; values greater than 2 provide robust evidence of causality.

### Protein domain annotation and 3-dimensional structure

To better understand the consequences of TR variation, we took advantage of the relationship between protein structure and function. We hypothesized that TRs that modify protein structure likely also influence protein function. Default parameters in AlphaFold2^23^ were applied with ColabFold v1.5.3^34^ to amino acid string sequences to predict the structure of a subset of proteins containing exonic TRs with fine-mapping probabilities >0.95. We predicted protein structure given the shortest and longest TR allele length alleles observed at least six times in the European ancestry population from UKB. Per-residue predicted local distance difference test (pLDDT) scores for expanded and contracted proteins were compared to the canonical sequence (accessed via UniProt^35^) to identify changes in local protein folding confidence. The pLDDT is a per-residue confidence metric that evaluates local distance differences of all atoms in a model, including stereochemical plausibility.^23^ As a measure of local model quality, a pLDDT value less than 50 is considered a strong predictor of structural disorder.^23^

### Mediation of psychiatric symptoms

We evaluated whether the TR-subcortical volume associations detected here reflected the mediating effects of subcortical volumes on psychiatric symptoms. From the UKB, we investigated the 7-item generalized anxiety disorder (GAD-7) questionnaire used to assess generalized anxiety disorder symptom severity, the 9-item patient health questionnaire (PHQ-9) used to assess depression symptom severity, and the 6-item posttraumatic stress disorder (PTSD) checklist (PCL-6) used to assess PTSD symptom severity. For GAD-7 and PHQ-9, participants were asked to respond with how often they have been bothered by the symptom/problem in the last two weeks. Participant responses range from “0” = “not at all” to “4” = “nearly every day”. For PCL-6, participants responded with how often they have been bothered by the symptom/problem in the past month. Participant responses range from “0” = “not at all” to “4” = “extremely”.

For each TR that was associated with subcortical volumes, we verified the TR was also associated with the GAD-7, PHQ-9, and PCL-6 questionnaire items using the same generalized linear models and covariates from the TR discovery phase. We considered any TR-psychiatric trait association with P<0.05 as suitable for inclusion in mediation analysis. We compared two models with the R package `mediation`^36^: (i) TR→subcortical volume (from our discovery phase) and (ii) TR + subcortical volume → psychiatric trait. Significant mediating effect of subcortical volume was determined if the average causal mediation effect (ACME) of subcortical volumeàpsychiatric trait effect was statistically different from null expectation (P<0.05).

## RESULTS

### Gene discovery per subcortical region

We associated 15,947 TRs from whole exome sequencing data with the volumes of seven subcortical brain regions. Per-trait lambda values ranged from 0.97 (pallidum) to 1.04 (accumbens) suggesting a lack of test statistic inflation due to sources other than polygenicity. After multiple testing correction (FDR< 5%; performed per subcortical region), we identified 19 significant TR-subcortical volume associations, of which, five had p-values less than a conventional GWAS threshold of P<5x10^-8^ (Figures 1A, S1, S2, and Table S1). These included intronic *ABLIM2*-[ACCC]_N_ and accumbens volume (β=6.53, se=1.16, P=1.92x10^-8^), *SPTBN4*-[GGGGC]_N_ and accumbens volume (β=4.37, se=0.77, P=1.50x10^-8^), and *ZMIZ2*-[GTGGG]_N_ and accumbens volume (β=8.79, se=1.51, P=6.25x10^-9^) and the 3’UTR repeat *NTN1*-[GCGG]_N_ and accumbens volume (β=5.93, se=1.03, P=8.16x10^-9^). The fifth genome-wide significant locus is an exonic repeat with an incredibly large effect on thalamus volume (*SLC39A4*-[CAG]_N_; β=-1599, se=286.7, P=2.42x10^-8^). The accumbens volume association analysis resulted in 13 study-wide associations (FDR<5% corresponding to P<5x10^-5^). These effects were concordant across the six other subcortical volumes (Figure 1B) with slopes ranging from 0.54 (P=9.94x10^-5^) in the pallidum to 2.98 (P=6.40x10^-29^) in the thalamus. These effects are reflected in the abundant nominally significant (P<0.05) overlap in observed effect sizes per TR in Figure 1A.

**Figure 1.**
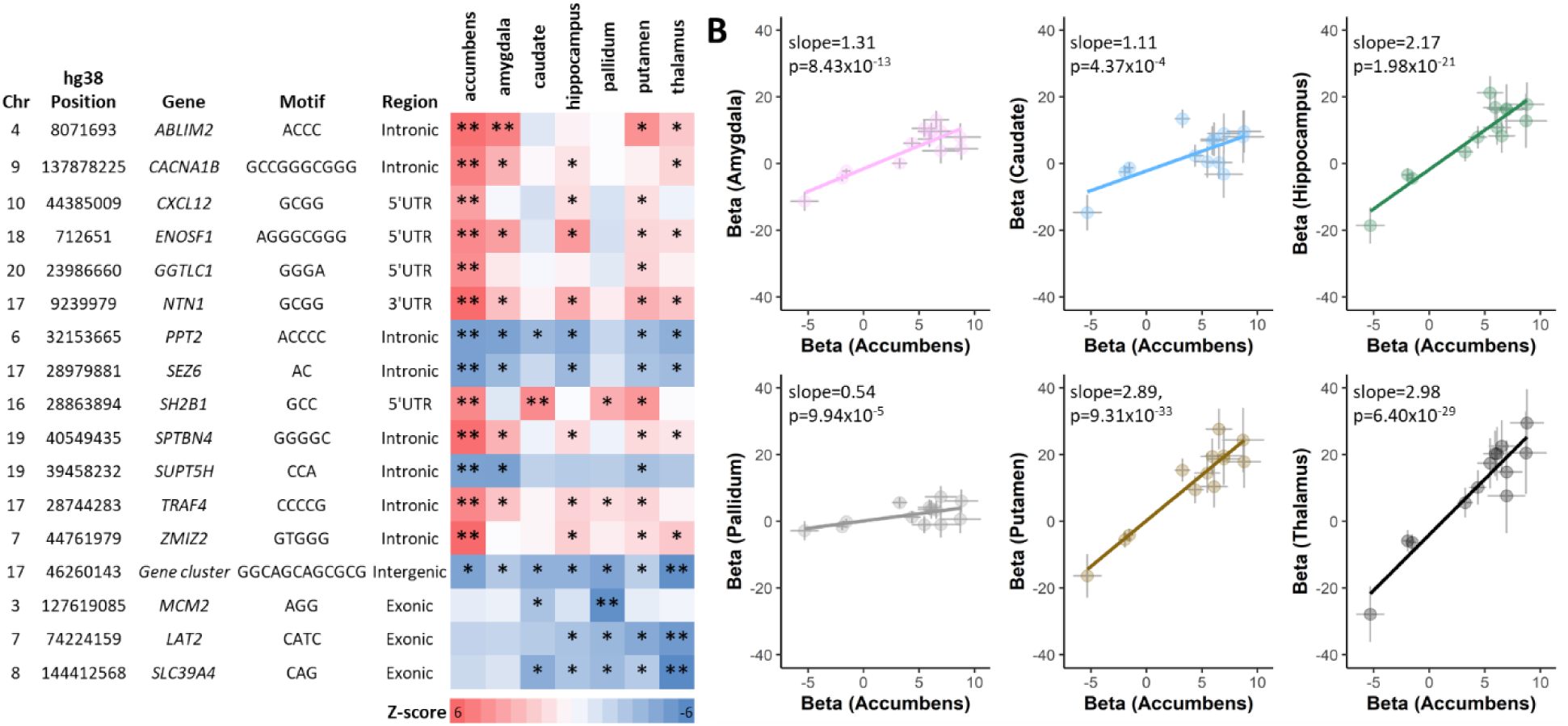
Locus discovery and tandem repeat annotation. (A) Eighteen trait-wide significant TR-subcortical volume association (FDR<5% denoted by **) with locus position and motif details. Nominally significant findings across subcortical regions are labeled with a single asterisk. (B) Prediction of non-accumbens TR effect sizes using study-wide significant TRs associated with the accumbens subcortical volume. Error bars around each data point reflect the standard error. Slopes of each line were estimated using no intercept and is weighted by the standard error of the y-axis subcortical volume observations.

The majority of genome-wide significant loci contained only a single causal variant. Among these, four genome-wide significant TRs had considerably high evidence of causality (log_10_BF > 2 and PIP > 0.98): *ABLIM2*-[ACCC]_N_, *NTN1*-[GCGG]_N_ and *SPTBN4*-[GGGGC]_N_ associated with accumbens volume and *SLC39A4*-[CAG]_N_ one associated with thalamus volume (Table S2). Notably, FINEMAP also prioritized a fifth putative causal variant in a locus with multiple associated TRs, as evidenced by the resolution of *SPTBN4*-[GGGGC]_N_ (PIP=0.987) over *SUPT5H*-[CCA]_N_ (PIP=0.013) as the causal variant in that locus.

### Cross-population replication

We sought to replicate results from European ancestry discovery samples were replicated in African (N=182), admixed American (N=53), Central/South Asian (N=349), East Asian (N=179), and Middle Eastern (N=58) ancestry populations. Despite reduced statistical power due to sample size, one association (*SH2B1*-[GCC]_N_) nominally associated with caudate volume in the Admixed American population: β=102.21, se=47.23, P=0.048 (Table S3). The AMR effect size for this relationship was not different (P=0.071) from the EUR effect size suggesting the large beta results from reduced statistical power rather than a truly larger effect on caudate volume in AMR participants.

### Gene Set Enrichment

Seven pathways were enriched for genes associated with subcortical volume using three pathway enrichment tools: g:Profiler, ShinyGO, and Enrichr (Figure 2A). Most pathways were specific to axon guidance and signaling and all were related to nervous system development. Four subcortical-volume associated genes were shared across several pathways: *ABLIM2*, *CXCL12*, *NTN1*, and *SPTBN4* (Figure 2B). The roles of *ABLIM2* (protein enabling actin filament binding), *NTN1* (signaling protein for axon guidance), and *CXCL12* (a chemokine) in the KEGG axon guidance pathway (P=7.43x10^-5^) are depicted in Figure S3, specifically in the context of axon outgrowth and repulsion. Tissue transcriptomic profile enrichment from GTEx v8 supports differential expression in the putamen basal ganglia (P=4.76x10^-5^), nucleus accumbens basal ganglia (P=1.72x10^-4^), and caudate basal ganglia (P=2.18x10^4^). However, up- or down-regulation of genes in these tissues were not identified (Figure S4 and Table S5).

**Figure 2.**
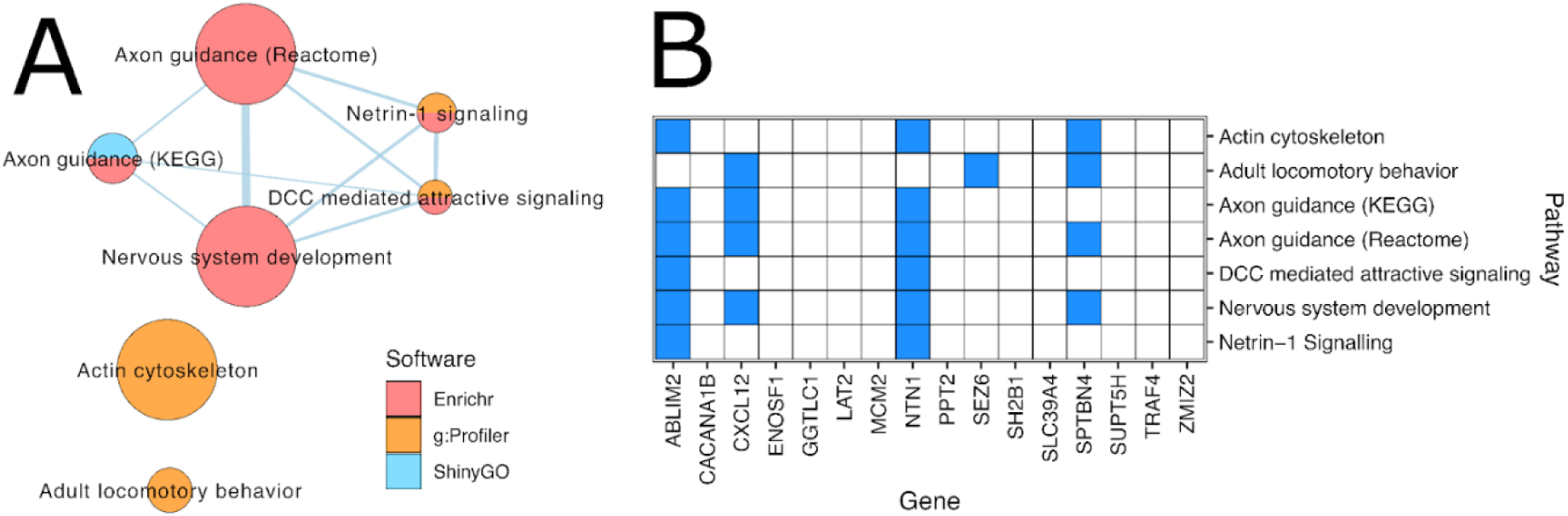
Pathway and tissue enrichment. A) Enrichment map depicting pathways significantly enriched for genes significantly associated with subcortical volume. Node size corresponds to gene set size and edge thickness corresponds to the number of shared genes between gene sets. Nodes are coloured by the enrichment software used to identify the pathways. (B) Plot showing significant genes and pathways. Genes that are part of the respective gene sets are coloured in blue.

### Gene expression and splicing effects

*SH2B1*-[GCC]_N_ was associated with increased splice quantity in 35 tissues, including anterior cingulate cortex (slope=0.212, P=3.79x10^-11^) and frontal cortex (Brodmann area 9; slope=0.145, P=5.28x10^-8^), and associated with decreased splice quantity in three tissues including cerebellar hemisphere (slope=-0.140, P=9.18x10^-7^) and nucleus accumbens basal ganglia (slope=-0.163, P=1.77x10^-13^; Figure 3, Table S5, and Table S6). *SUPT5H*-[CCA] was consistently associated with decreased splice quantity across 12 tissues including spinal cord (cervical C1; slope=-0.675, P=2.49x10^-7^) and EBV transformed lymphocytes (slope=-0.578, P=1.52x10^-6^).

**Figure 3.**
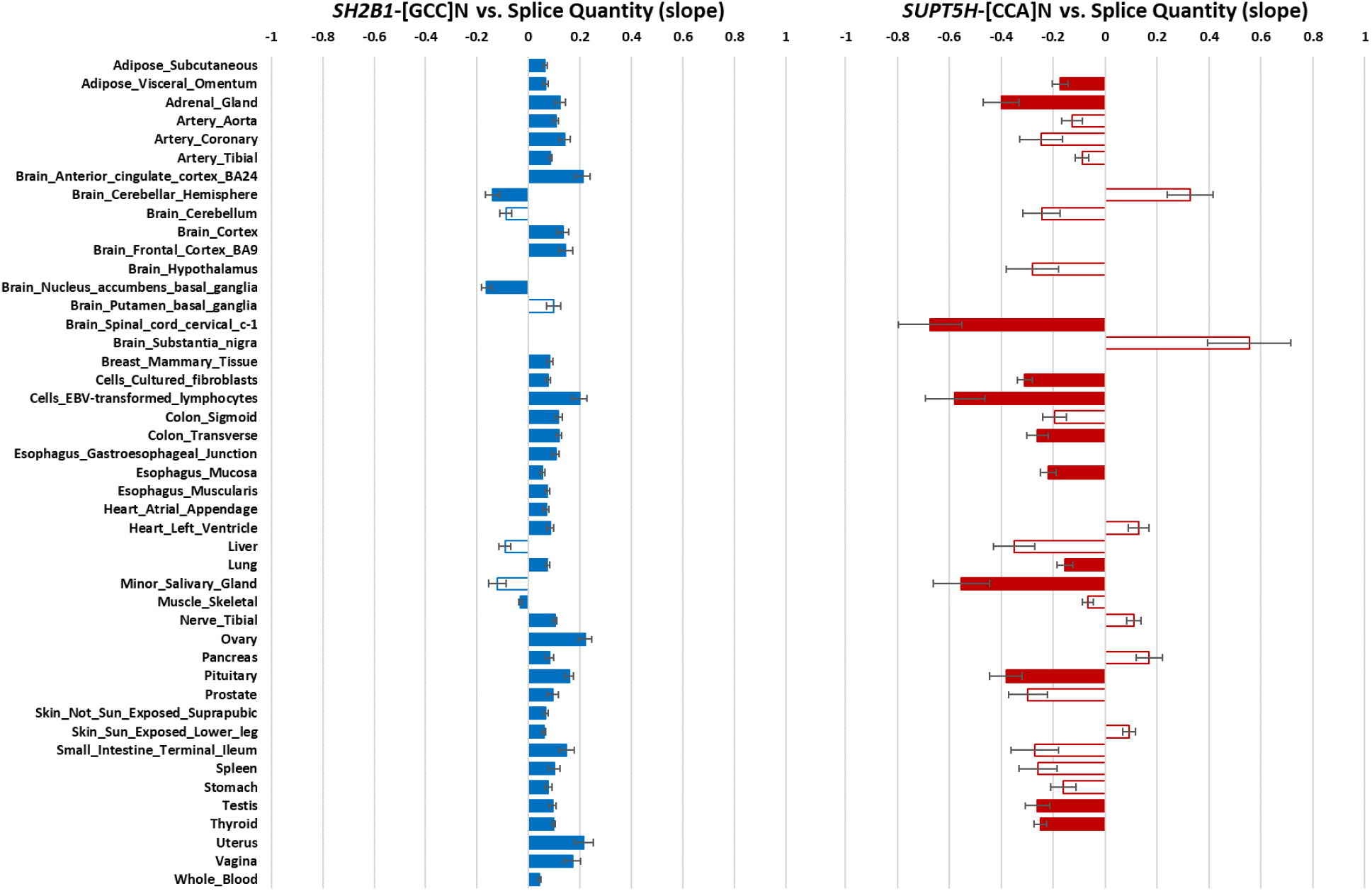
Splicing effects of subcortical volume associated TRs based on findings from Hamanaka, et al.^31^ using GTEx v8 (49 tissues in N=652 unrelated donors). Solid bars indicate significant spl-TR slopes with error bars reflecting the standard error for each slope.

### Prioritization of *NTN1*-[GCGG]_N_

The discovered TRs were subjected to several layers of analysis to describe their effect on the associated subcortical region. We describe two noteworthy examples of variant prioritization using these annotation procedures: (i) *NTN1*-[GCGG]_N_ as an example TR with a wide range of common alleles and (ii) *LAT2*-[CATC]_N_ as an example TR with a narrow range of rare alleles.

At *NTN1*-[GCGG]_N_, there was a 59mm^3^ difference in accumbens volume between the smallest and largest repeat burden (Cohen’s *d*=0.279, P=3.51x10^-4^; Figure 4A). Relative to the modal genotype ([GCGG]_3_|[GCGG]_3_), each change in repeat unit length had a minor effect on organism fitness (*s*=0.005, P=0.041; Table S8). Appreciating the relationship between subcortical volumes and psychiatric symptoms, we tested whether *NTN1*-[GCGG]_N_ had either mediated or independent influence on symptoms of major depression, generalized anxiety, and posttraumatic stress. In addition to its effect on accumbens volume, *NTN1*-[GCGG]_N_ was associated with the PTSD symptom “felt very upset when reminded of stressful experience” (UKB Field ID 20498, β=0.012, se=0.005, P=0.010). A small portion of this effect was mediated by the relationship between accumbens volume and upset feelings (Figure 4B and Table S9, β=3.85x10^-4^, se=1.95x10^-4^, P=0.048) such that *NTN1*-[GCGG]_N_ remained associated with PTSD symptoms after adjusting for accumbens volume (β=0.012, se=0.005, P=0.011). In addition to accumbens volume, NTN1 has several pleiotropic effects with neurological (1.57-fold enrichment, P=2.65x10^-5^) and dermatological (4.16-fold enrichment, P=3.02x10^-6^) traits domains (Figure 4C and Table S10).

**Figure 4.**
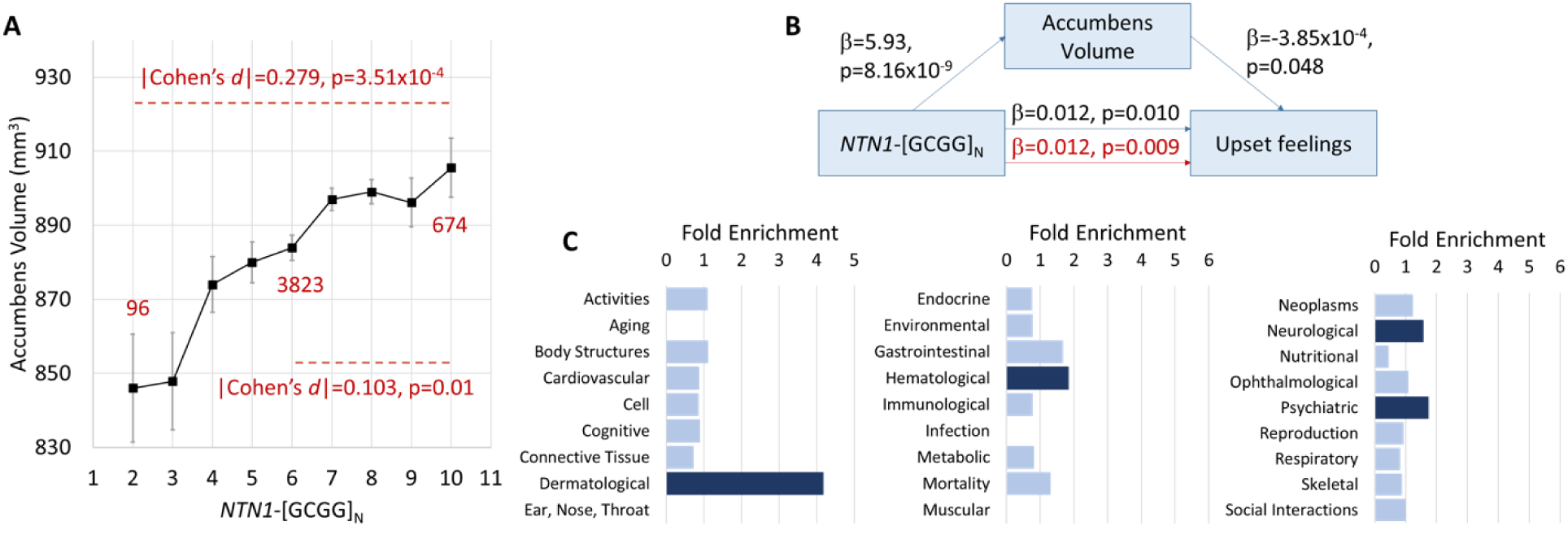
Example of a subcortical volume associated tandem repeat with a wide range of common alleles. (A) Association between locus level repeat burden at *NTN1*-[GCGG]_N_ and accumbens volume. Sample sizes are shown for the modal repeat burden (6, genotype ([GCGG]_3_ | [GCGG]_3_)) and the largest expansions and contractions seen in the UK Biobank European ancestry participants. Cohen’s *d* estimates were calculated between the two extreme ends of the repeat burden distribution as well as the modal allele versus the largest expansion. (B) *NTN1*-[GCGG]_N_ effects the posttraumatic stress symptom “upset feelings” independently of accumbens volume estimated by mediation analysis (red text denotes the direct effect after controlling for accumbens volume). Mediation analysis results across a variety of major depression, generalized anxiety, and posttraumatic stress symptoms are provided in Table S9. (C) Phenome-wide association study of *NTN1* in the GWAS Atlas (dark bars indicate p < 0.05). All enrichments and P-values (C) are provided in Table S10.

### Prioritization of *LAT2*-[CATC]_N_

At *LAT2*-[CATC]_N_, there was a 2.9cm^3^ difference in thalamus volume between the extreme ends of TR-burden (Cohen’s *d*=0.800, P=0.041; Figure 5A). Relative to the modal genotype ([CATC]_3_|[ CATC]_3_), each change in repeat unit length had a large effect on organism fitness (*s*=0.823, P=5.49x10^-5^; Table S8) consistent with the relative rarity of these repeat length burdens and the large effect conferred by expansion or contraction. This tetranucleotide repeat found from LAT2/NTAL amino acids 197-200 is part of a disordered region of the resulting protein containing a single helical structure. Despite having little influence on protein structural properties among the 196 unaltered amino acids (mean ΔpLDDT_canonical_=55.03±14.1; ΔpLDDT_expansion_=55.56±13.4, P_diff_=0.978; ΔpLDDT_contraction_=46.78±15.6, P_diff_=0.695), the expanded and contracted forms of *LAT2*-[CATC]_N_ lead to frameshift mutations that disrupt protein folding and prevent appropriate stop codon placement (Figure 5B). In addition to its effect on thalamus volume, *LAT2*-[CATC]_N_ was associated with four major depression symptoms, two generalized anxiety symptoms, and one posttraumatic stress symptom (Figure 5C and Table S9). The largest of these effects was with the PTSD symptom “felt very upset when reminded of stressful experience” (UKB Field ID 20498, β=0.426, se=0.138, P=0.002, Figure 5C, Figure S5, and Table S9).

**Figure 5.**
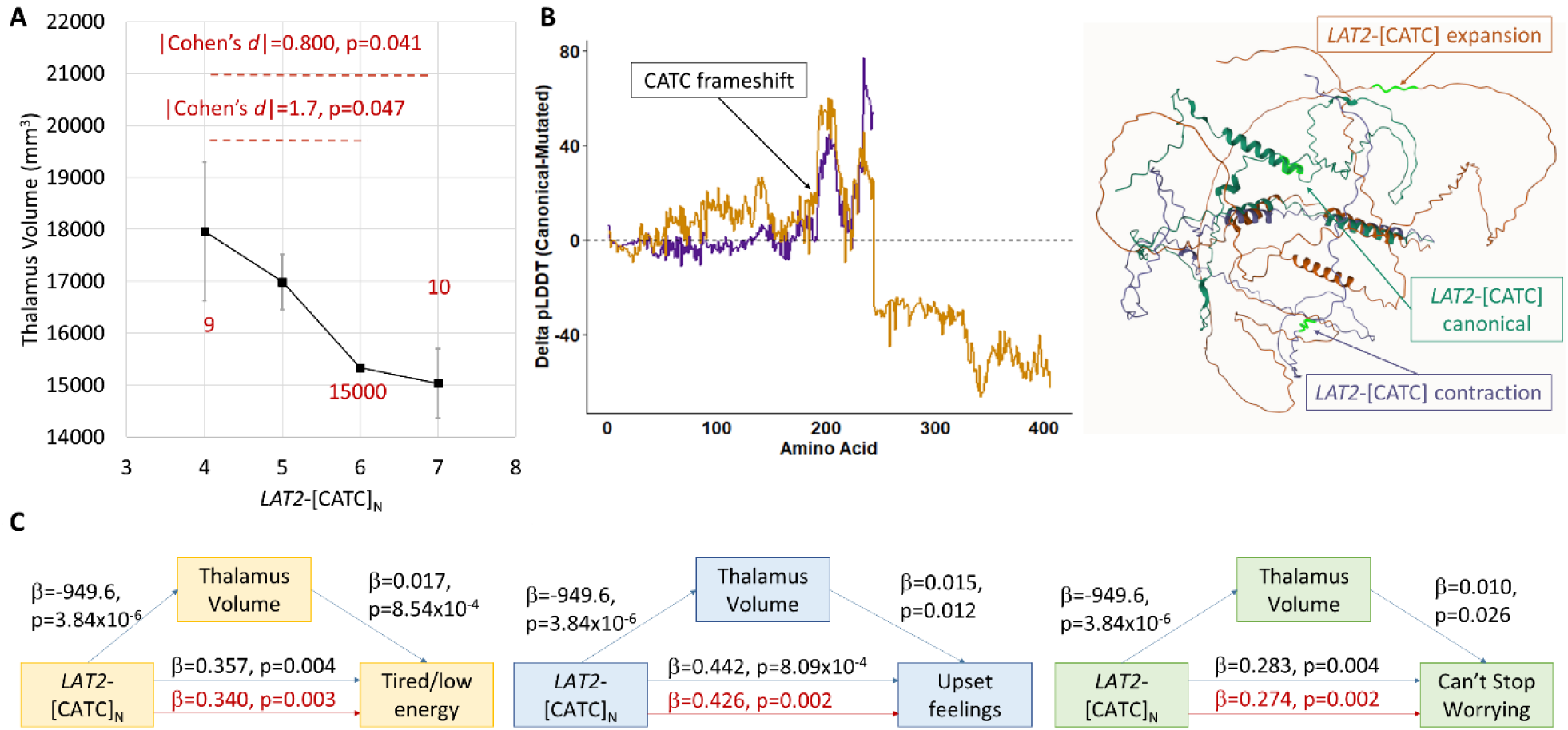
Example of a subcortical volume associated tandem repeat with a narrow range of rare alleles. (A) Association between locus level repeat burden at *LAT2*-[CATC]_N_ and thalamus volume. Sample sizes are shown for the modal repeat burden (6, genotype [CATC]_3_ | [CATC]_3_) and the largest expansions and contractions seen in the UK Biobank European ancestry participants. (B) Change in per-residue folding confidence (ΔpLDDT) between canonical *LAT2* and contracted (purple) and expanded (orange) forms of *LAT2*-[CATC]_N_. Folded proteins were superimposed to show abnormal structure resulting from expansion and contraction of *LAT2*-[CATC]_N_. (C) *LAT2*-[CATC]_N_ effects the major depression (yellow), generalized anxiety (green), and posttraumatic stress symptoms (blue) independently of thalamus volume (red text denotes the direct effect after controlling for thalamus volume). Mediation analysis results across a variety of major depression, generalized anxiety, and posttraumatic stress symptoms are provided in Figure S5 and Table S9.

### Other Notable Prioritized TRs

Two other TRs associated with subcortical volumes were localized to exonic regions of their respective genes. The *MCM2*-[AGG]_N_ repeat encodes a stretch of glutamate amino acid residues in a highly disordered region of MCM2 (minichromosome maintenance complex component 2). Contraction of this repeat unit conferred a large increase in pallidum volume (1.56cm^3^) between the modal genotype and contracted form (Cohen’s *d*=3.39, P=0.017, Table S7 and Figure S11). Despite being exonic, contraction of this repeat unit had no significant influence on protein folding (Figure S12). The largest change in local per-residue folding confidence was found just downstream of the *MCM2*-[AGG]_N_ repeat at lysine-696 (ΔpLDDT=-11.4) in favor of the contracted form of *MCM2*-[AGG]_N_. This minor effect was supported by lack of an effect of this TR on organism fitness (Table S8). However, *MCM2*-[AGG]_N_ had a relatively large effect on the major depression symptom “trouble falling asleep of staying asleep, sleeping too much” (UKB Field ID 20517, β=0.311, se=0.151, P=0.044, Table S9).

The *SLC39A4*-[CAG]_N_ repeat encodes a stretch of lysine amino acid residues in an unstructured region of zinc transporter ZIP4. Contraction of this repeat unit conferred a large increase in thalamus volume (5.03cm^3^) between the modal genotype and contracted form (Cohen’s *d*=3.40, P=5.29x10^-4^, Table S7 and Figure S11). Despite being exonic, contraction of this repeat unit had no significant influence on protein folding (Figure S12). The largest change in local per-residue folding confidence was found in the last amino acid of SLC39A4 (ΔpLDDT=16.28) in favor of the canonical form of *SLC39A4*-[CAG]_N_. This benign repeat contraction had no evidence of effect on organism fitness (Table S8) and, despite enrichment of neurological pleiotropic effects, had no association with symptoms of major depression, generalized anxiety, or posttraumatic stress (Tables S9 and S10).

## DISCUSSION

Prior work has investigated TRs associated with a similar phenotype, grey matter volume,^37^ using TR genotypes derived from ExpansionHunter^38^ and reported an enrichment of caudate- and pallidum-associated loci. This work used 74 finer-grained FAST magnetic resonance image-derived phenotypes and identified 83 TRs within a gene boundary (though not necessarily exonic) and 63 TRs in intergenic regions. Several genes identified in this TR-based analysis did not have any significant SNP associations in GWAS of the same image-derived phenotypes, supporting that TR investigations can uncover novel insights into the genetics of brain-related outcomes. The subcortical brain structures play key roles in cognitive and emotional functions and their volumetric variation is often implicated in psychiatric symptoms and diagnoses.^1,39^ We conducted an exome-wide association study of seven subcortical brain volumes using TR variation and identified 17 TRs with compelling evidence for a role in brain morphology. Many of the loci uncovered here exhibited high fine-mapping probabilities relative to surrounding variation, influence splicing, have independent effects on psychiatric symptoms, and even modify protein 3-dimensional structure.

The most significantly associated variant uncovered was found in *NTN1*.^40^ *NTN1* encodes netrin-1, an evolutionarily conserved secreted protein that plays a crucial role in axon guidance during embryonic development.^41^ Netrin-1 contributes to various physiological processes, including angiogenesis, cell survival, and synaptic plasticity. Our study identified that a 3’UTR motif *NTN1*-[GCGG]_N_ associated with accumbens volume. This localization to the 3’UTR suggests a regulatory role. However, this TR was not associated with cis-gene expression or cis-splicing effects in the GTEx database. *NTN1* does play a major role in the developing spinal cord based on the balanced expression of coding DNA sequence and 3’UTR sequence.^41^ Netrin-1 activates a translation-dependent increase in β-actin leading to excess axonal growth cones and highly branched neurons.^42,43^ The netrin-1 signaling pathway, including its effect on β- actin and its receptor DCC, was studied extensively in the context of depression and schizophrenia.^44,45^ Our study highlights a direct role of *NTN1*-[GCGG]_N_ on both accumbens volume and depression symptoms. For these reasons, further research may investigate whether the discovered TR directly contributes to mood disorder vulnerability via the ability of netrin-1 to increase β-actin levels and/or influence expression of a gene(s) from a related pathway (i.e., in a trans-regulatory manner).

The largest effect size TR that we identified was *LAT2*-[CATC]_N_ in relation to thalamus volume and independently influencing seven psychiatric symptoms across the internalizing spectrum. *LAT2* encodes the Linker For Activation Of T Cells Family Member 2 and, along with LAT1, is a primary transporter of tryptophan and 5-hydroxytryptophan.^46^ The detected *LAT2* TR is localized to a four amino acid region directly flanked by two post-translationally modified residues. In its expanded and contracted form, *LAT2*-[CATC]_N_ causes a frameshift mutation that largely disrupts protein folding by disrupting the position of the normal stop codon. Consequently, translation machinery may polymerize up to 157 additional residues following the addition of a single repeat motif (*LAT2*-[CATC]_4_). LAT2 is a target for the treatment of depression using the selective serotonin reuptake inhibitor fluoxetine by controlling the availability of tryptophan in the brain.^46^ Reduced LAT2 abundance caused by frameshift mutation we discovered may contribute to treatment-resistant depression.^46^ Furthermore, *LAT2* is located within the critical region associated with Williams-Beuren syndrome (WBS). WBS is caused by a hemizygous deletion of a 1.5-1.8Mb region of chromosome 7 that spans up to 28 genes.^47^ Though found within that critical region, *LAT2* is not always deleted among cases who carry the smaller 1.5Mb deletion.^48,49^ WBS has a heterogeneous set of symptoms, including cardiovascular, cognitive, renal, dental, and gastrointestinal anomalies, and various neurodevelopmental outcomes.^47^ Given the rarity of this TR expansion and contraction observed among UK Biobank participants, and the documented relationship of *LAT2* with WBS and autism spectrum disorder^50^, we hypothesize that the *LAT2*-[CATC]_N_ expanded and contracted states reduce LAT2 protein concentration, contributing to an array of neuropsychiatric outcomes.

We identified two additional exonic TRs with high fine-mapping probabilities suggesting their role as a putative causal variation for pallidum (*MCM2*-[AGG]_N_) and thalamus (*SLC39A4*-[CAG]_N_) volume. Though exonic, the effects of the expanded and contracted forms of the detect repeat motifs were relatively benign in terms of protein folding. MCM2 encodes minichromosome maintenance complex component 2, which facilitates DNA replication initiation.^51^ In the adult brain, MCM2 is essential for maturation of dentate gyrus cells via their transition from neural progenitor cell to neuroblast.^52^ This transition eventually produces mature neurons, primarily in the hippocampus, that project through the pallidum to the thalamus. Despite this process being necessary for normal learning and memory, our study showed no relationship between *MCM2*-[AGG]_N_ and internalizing symptoms suggesting the effect on learning and memory is strictly through its effect on neurogenesis rather than other routes to influence psychopathology. *SLC39A4* encodes zinc transporter ZIP4. The role of zinc transport in psychiatry largely reflects a contribution to depression and psychosis via a critical modulatory role in neurotransmission.^53^

To prioritize how TRs might influence subcortical volume, we investigated both TR-associated gene expression and TR-associated splicing. *SH2B1*-[GCC]_N_ was generally associated with increased splicing across tissues. This is consistent with *SH2B1* having four common alternatively spliced isoforms. SH2B1α and SH2B1δ isoforms appear to be mostly brain specific.^54^ *SH2B1* was detected by lentiviral massively parallel reporter assay as a differentially activated autism spectrum disorder associated variant with effects on transcription factor binding and chromatin accessibility patterns specific to radial glia.^55^ The differential activation of this region of the genome may be tied to *SH2B1* splicing activity in the cerebellar hemisphere and the nucleus accumbens. *SUPT5H*-[CCA]_N_ was ubiquitously associated with reduced splicing in a relatively sparse collection of GTEx tissues and reflects a mix of brain and peripheral tissue. *SUPT5H* protein product is a post-transcriptional silencing factor and when knocked down together with SUPT4H1 reduces the accumulation of poly(GP) protein without large changes in other transcripts.^56^ While this has been proposed as one possible therapeutic for repeat-associated *C9ORF72* neurodegenerative disorders, the deleterious consequences due to *SUPT5H* influence on other genes are still unclear.^57^

Our study provides significant insights into the influence of repeat variation on subcortical volumes and psychiatric symptoms. However, it is important to note certain limitations that may restrict the scope of our findings. First, because we focused on common exonic repeats, we did not assess non-coding and rare TRs associated with subcortical volumes. Second, our attempt at cross-ancestry replication suffered from lack of statistical power. The relatively small sizes of diverse ancestry groups restricted our ability to replicate repeat-associated effects. Therefore, an extensive exploration across diverse ancestries is imperative to reveal population-specific nuances and enhance the overall robustness of our findings. Third, we chose to sum the left and right hemispheres of the each subcortical region and the heterogeneity associated with the summed phenotype may mask TR-associations relevant for hemispheric hypotheses. Finally, our use of the GangSTR approach leads to reduced accuracy of AC/TG dinucleotide repeat motifs. These are scarcely represented in our data yet should be interpreted with their elevated error rate in mind. Work is currently ongoing to harmonize genotypes across TR callers to help reduce error rates in contexts where one caller performs poorly relative to others.^58^

Our findings pave the way for a novel approach to investigating the influence of genetic factors on brain biology. We, and others,^15–17,59^ have demonstrated the power of TR-wide association studies to identify significant genetic effects independent of those discovered by SNP-focused GWAS. In particular, this findings pave the way for a novel approach to investigating the work expands our understanding of the genetic architecture of subcortical volumes, underscoring the importance of considering repetitive elements in the broader landscape of brain genetics.

## ACKNOWLEDGEMENTS

This research was performed using the UKB Resource under the application 58146. This project is partially supported by funding from the University of Toronto Data Sciences Institute (FRW and EJP) and the University of Toronto McLaughlin Centre (to FRW). RP acknowledges support from NIH (RF1 MH132337 and R33 DA047527) and One Mind.

## AUTHOR CONTRIBUTIONS

Conceptualization: FRW, PH; Methodology: MAP, YS, PH, FRW; Investigation/Analysis: MAP, YS, CLA, NA, AMC, FRW; Visualization: MAP, NA, FRW; Supervision: FRW; Writing – original draft: MAP, YS, FRW; Writing – review and editing: MAP, YS, CLA, NA, AMC, EJP, RP, PH, FRW; Study-specific funding acquisition: FRW, EJP.

## DATA AVAILABILITY

All subcortical volume summary association statistics are available for download at 10.5281/zenodo.10157819. All other data supporting the conclusions of this work are provided as supplementary material. This study used UK Biobank which data can be accessed by researchers who submit and received approval of a study protocol (https://www.ukbiobank.ac.uk/enable-your-research/apply-for-access).

## SUPPLEMENTARY INFORMATION

### FIGURES

**Figure S1.**
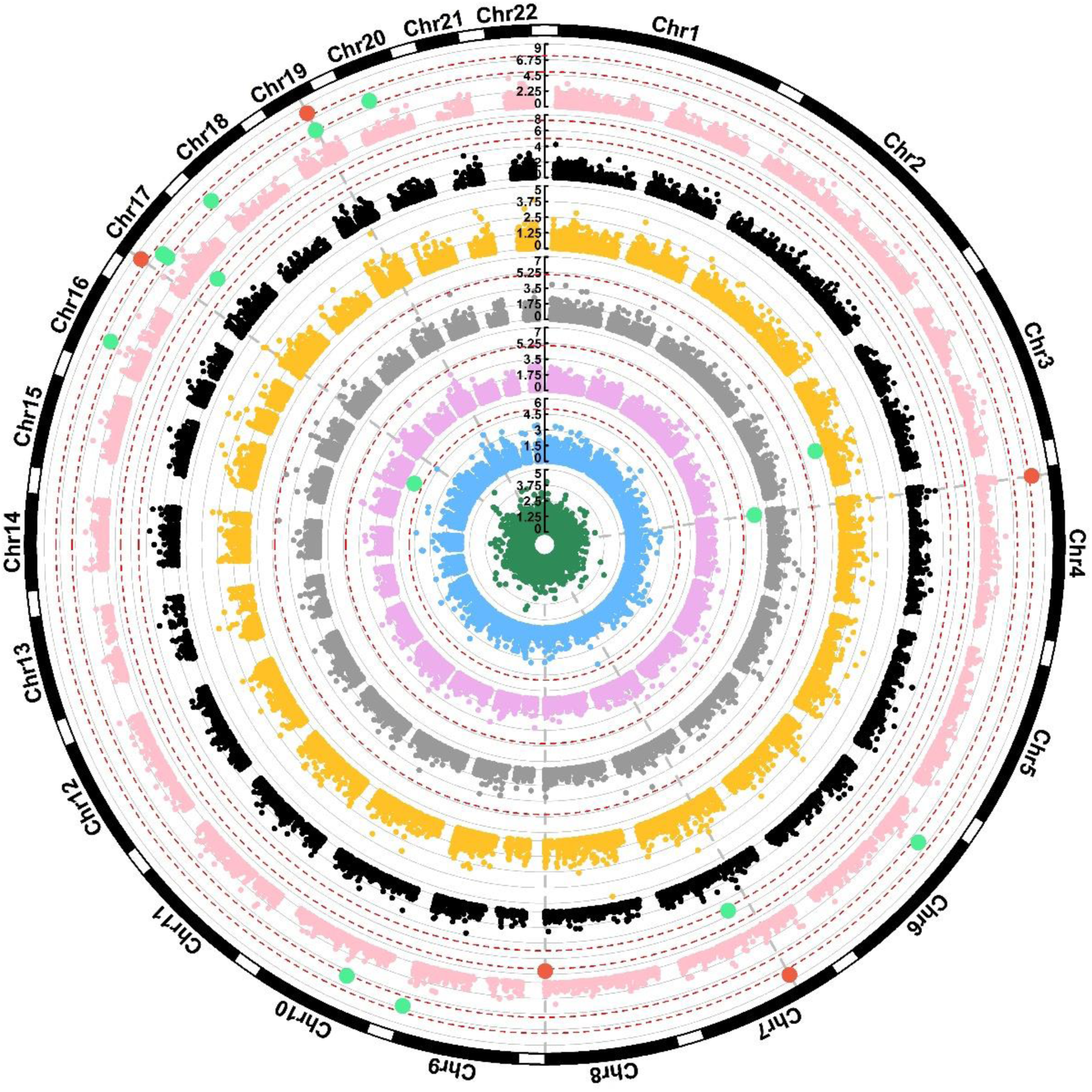
Circular Manhattan plot of tandem repeat associations with subcortical volumes. Subcortical volumes are ordered as hippocampus (green inner-most circle), caudate (blue), amygdala (plum), pallidum (grey), putamen (yellow), thalamus (black), accumbens (pink outer-most circle). Green data points survived a suggestive trait-wide false discovery rate of 5% and red data points survived a traditional genome-wide p-value threshold (P<5x10^-8^).

**Figure S2.**
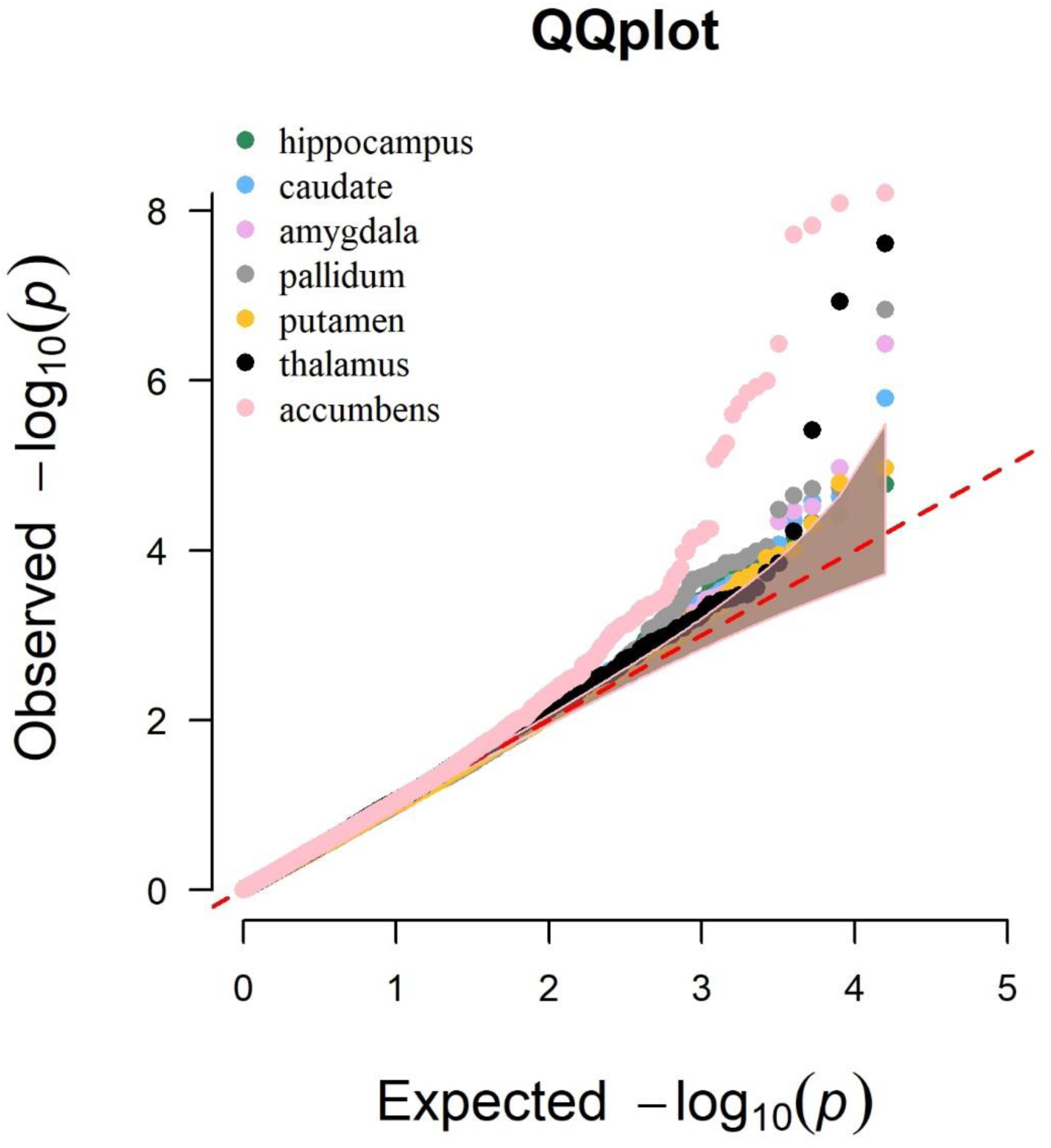
Quantile-quantile plots of the seven subcortical volume association studies. Lambda values, reflecting the distribution of p-values per association study compared to a normal distribution, ranged from 0.97 (pallidum) to 1.04 (accumbens).

**Figure S3.**
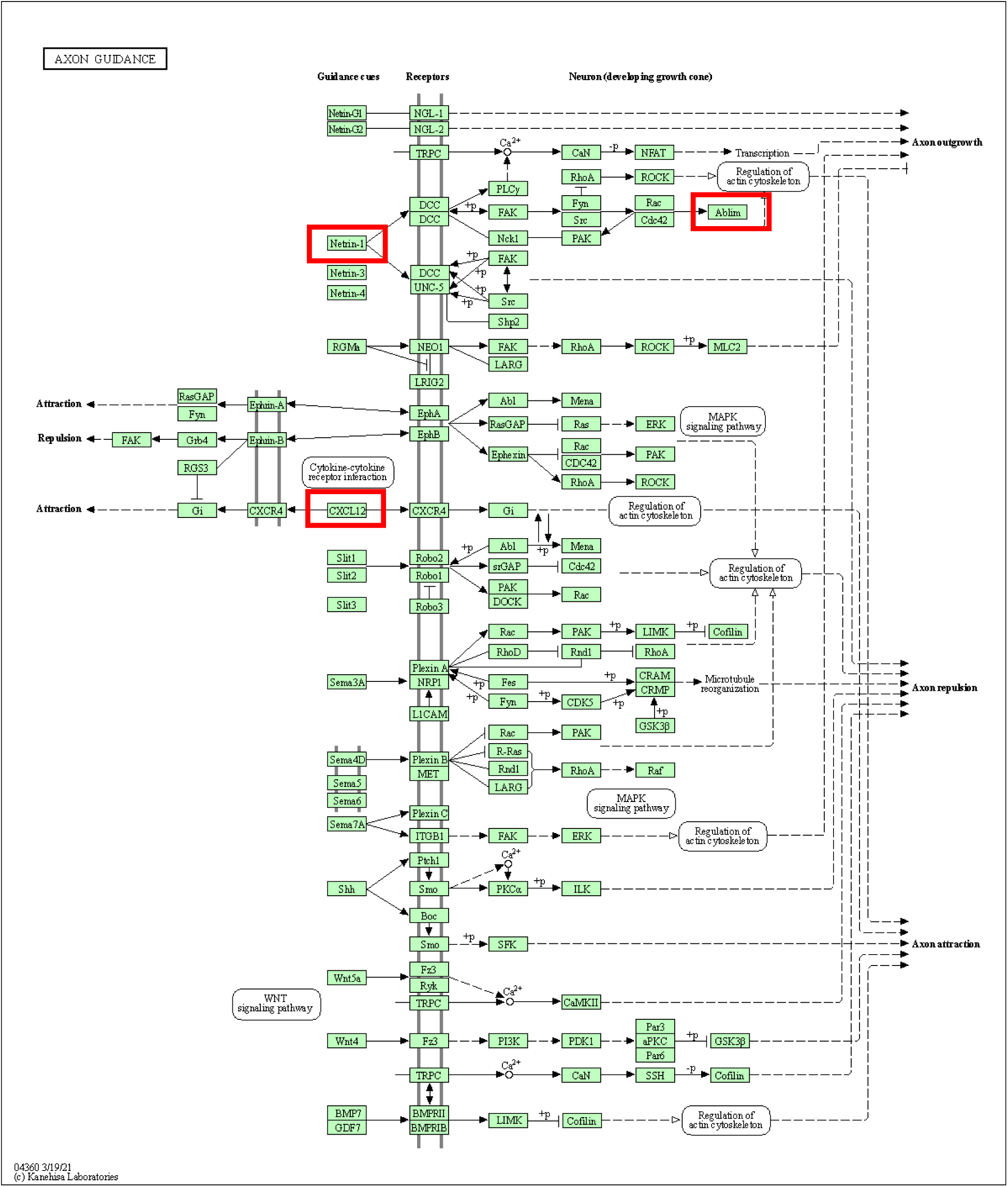
KEGG pathway for Axon Guidance with the three subcortical volume associated proteins highlighted in red.

**Figure S4.**
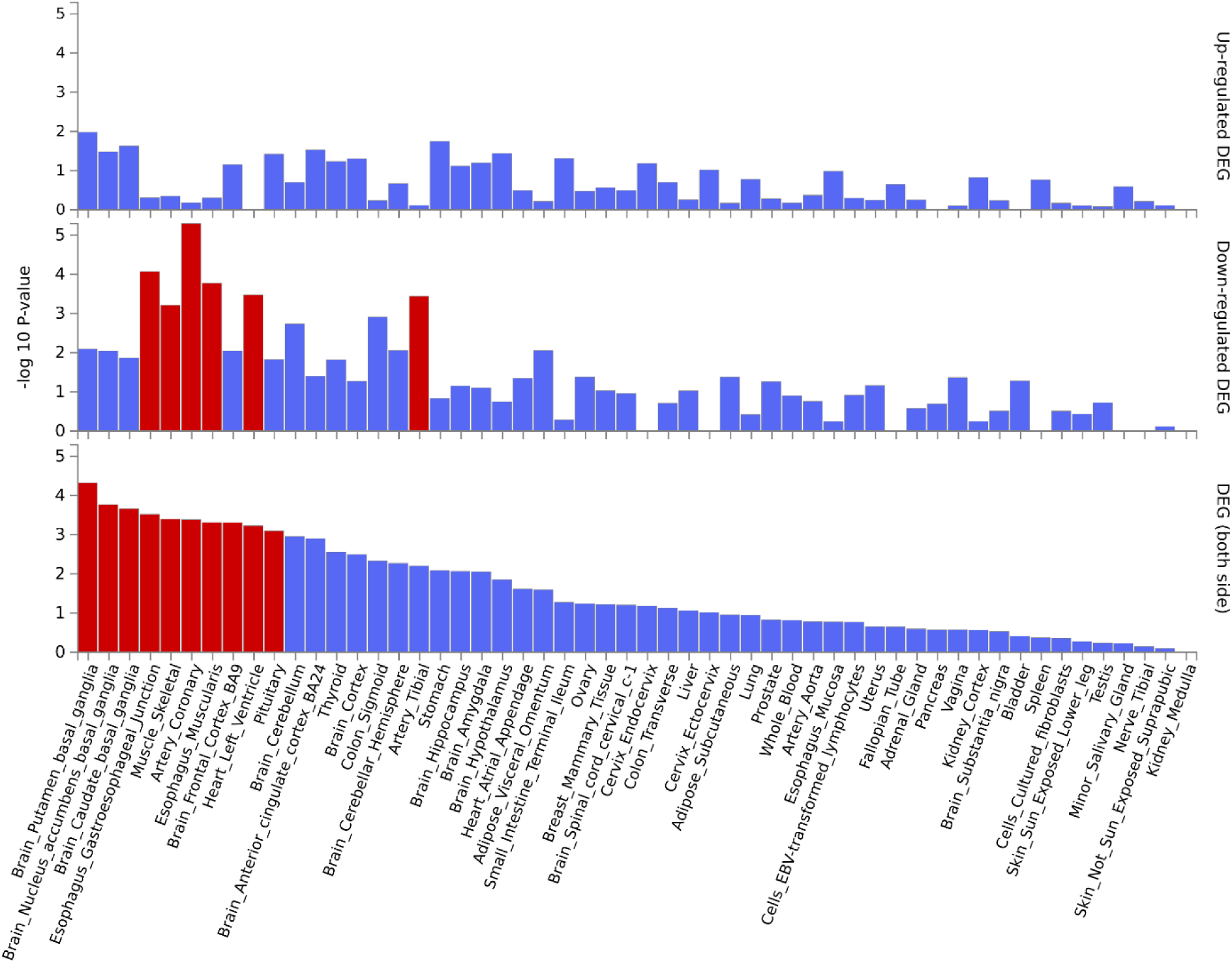
Tissue enrichment results from FUMA using the 30 general GTEx v8 tissues. Overlapping genes, enrichment p-values, and adjusted p-values are reported in Table S4.

**Figure S5.**
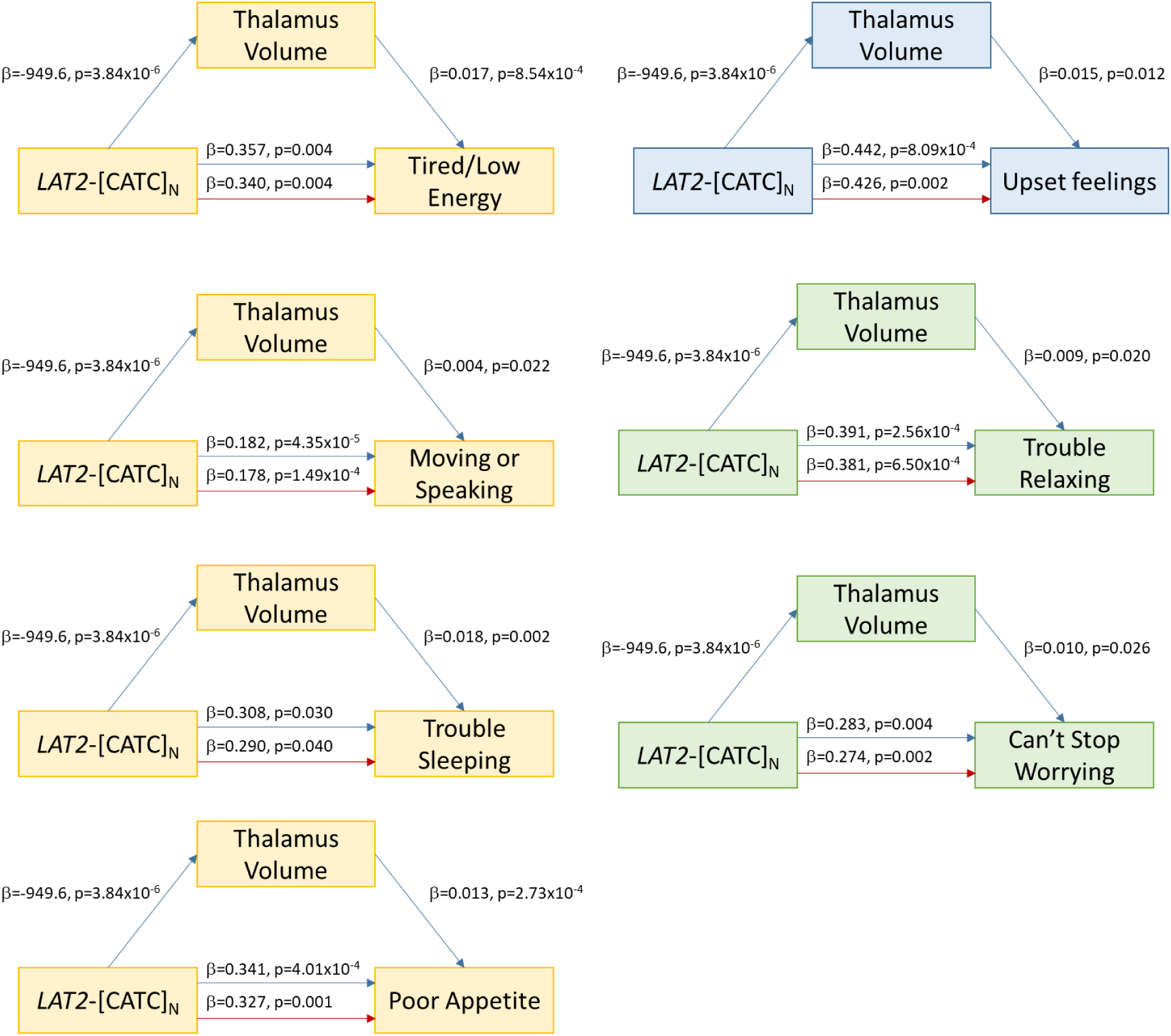
Significant independent effects of *LAT2*-[CATG]_N_ on depression (yellow), generalized anxiety (green), and posttraumatic stress (blue) symptoms independent of thalamus volume. Red text denotes the direct effect (ADE) after controlling for the relationship between thalamus volume and each psychiatric symptom. “Tired/Low Energy” = UKB Field ID 20519, “Moving or Speaking” = UKB Field ID 20518, “Trouble Sleeping” = UKB Field ID 20517, “Poor Appetite” = UKB Field ID 20511, “Upset Feelings” = UKB Field ID 20498, “Trouble Relating” = UKB Field ID 20515, “Can’t Stop Worrying” = UKB Field ID 20509. Refer to Table S9 for additional details for each mediation analysis.

**Figure S6.**
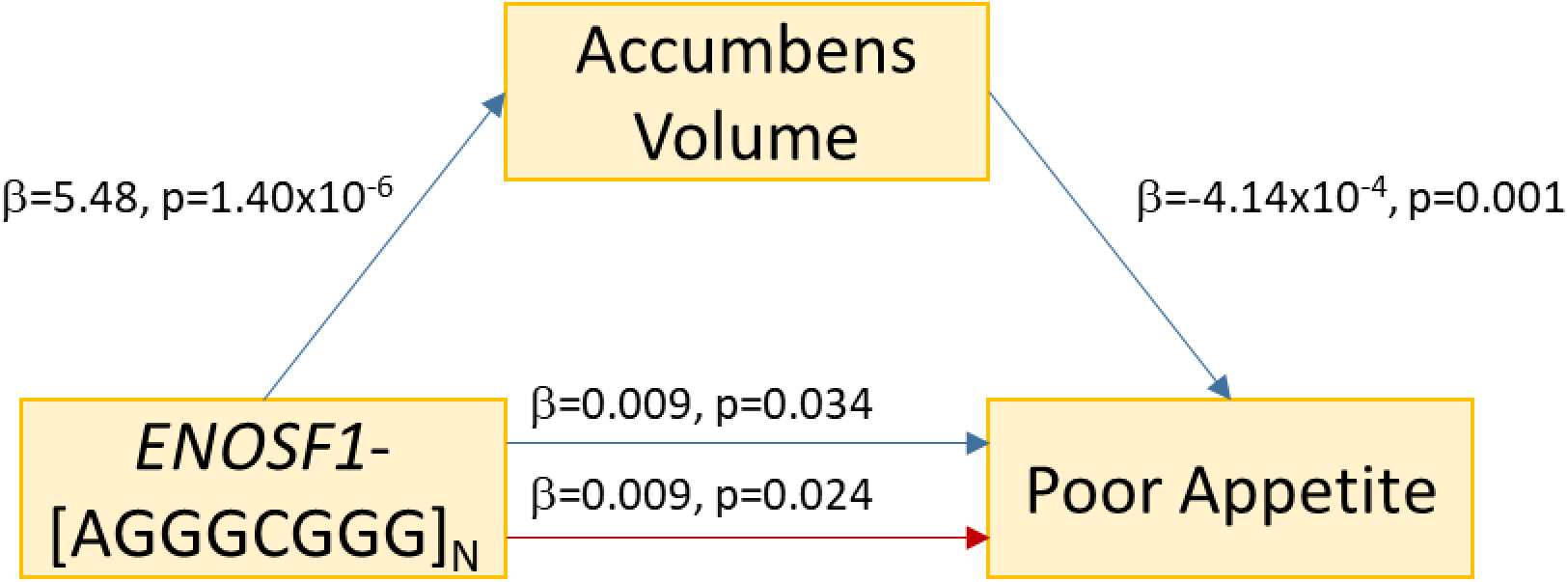
Significant independent effects of *ENOSF1*-[AGGGCGGG]_N_ on depression symptoms independent of accumbens volume. Red text denotes the direct effect (ADE) after controlling for the relationship between accumbens volume and poor appetite (UKB Field ID 20511). Refer to Table S9 for additional details for each mediation analysis.

**Figure S7.**
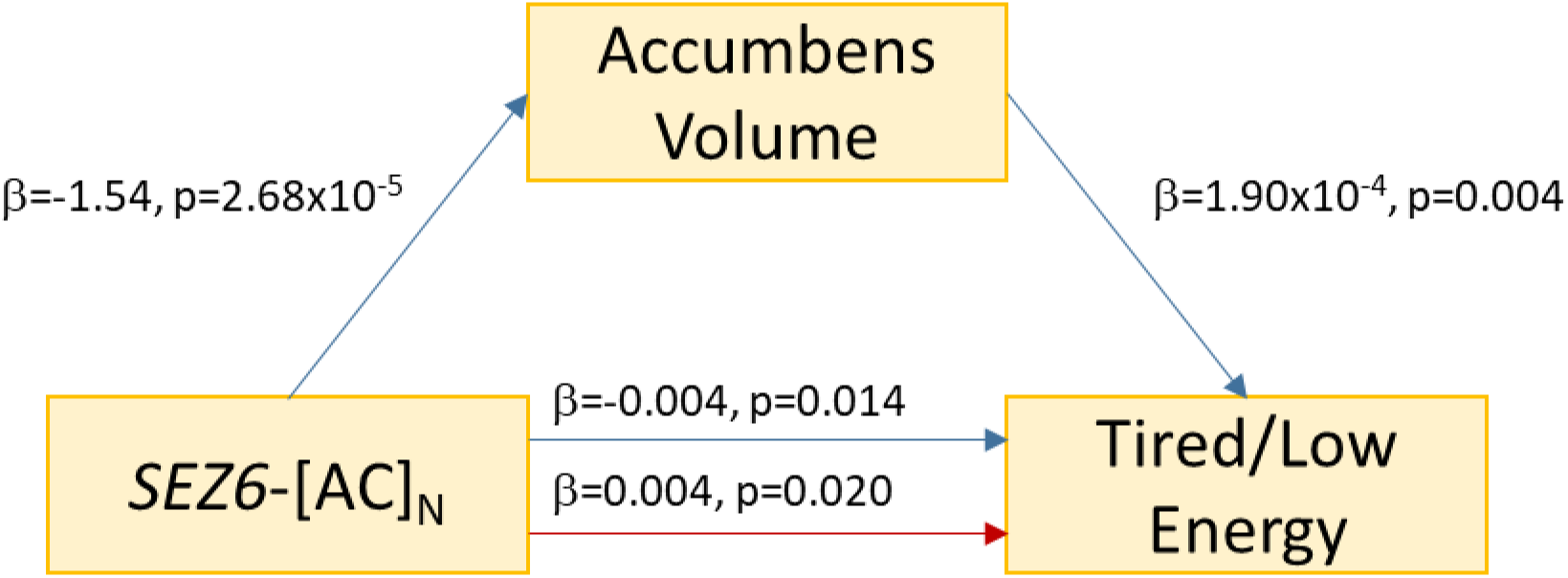
Significant independent effects of *SEZ6*-[AC]_N_ on depression symptoms independent of accumbens volume. Red text denotes the direct effect (ADE) after controlling for the relationship between accumbens volume and tired/low energy (UKB Field ID 20519). Refer to Table S9 for additional details for each mediation analysis.

**Figure S8.**
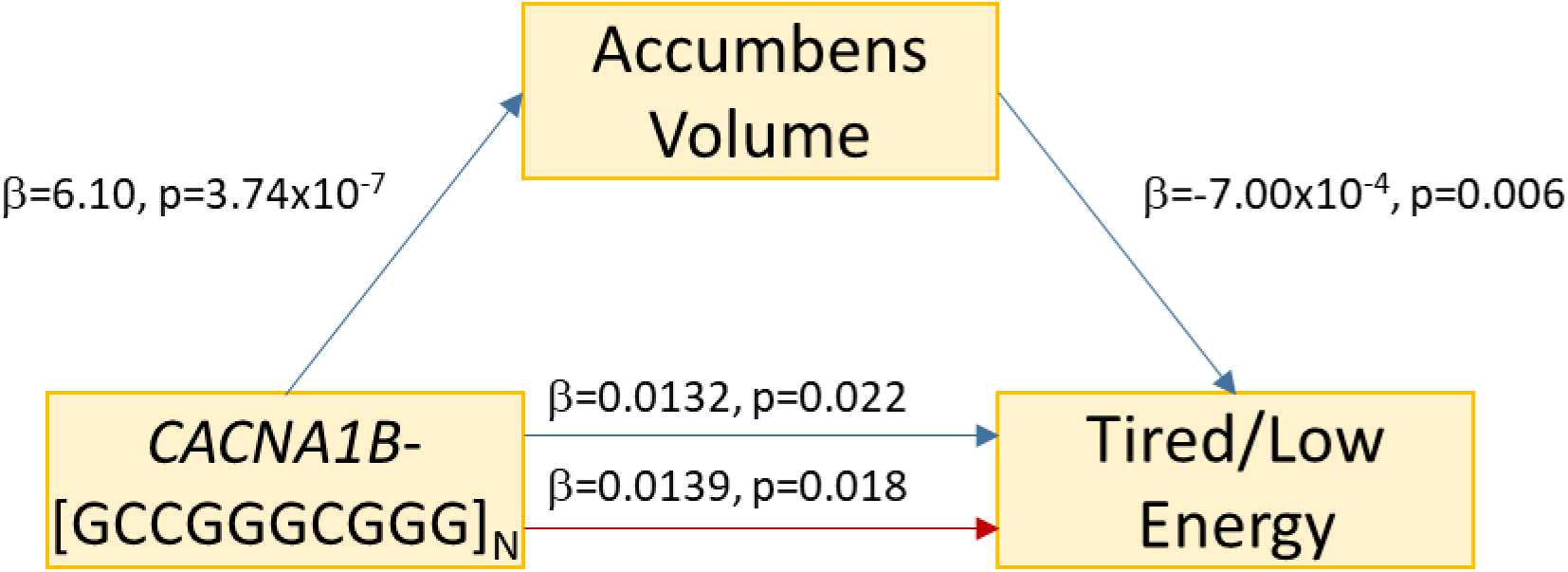
Significant independent effects of *CACNA1B*-[GCCGGGCGGG]_N_ on depression symptoms independent of accumbens volume. Red text denotes the direct effect (ADE) after controlling for the relationship between accumbens volume and tired/low energy (UKB Field ID 20519). Refer to Table S9 for additional details for each mediation analysis.

**Figure S9.**
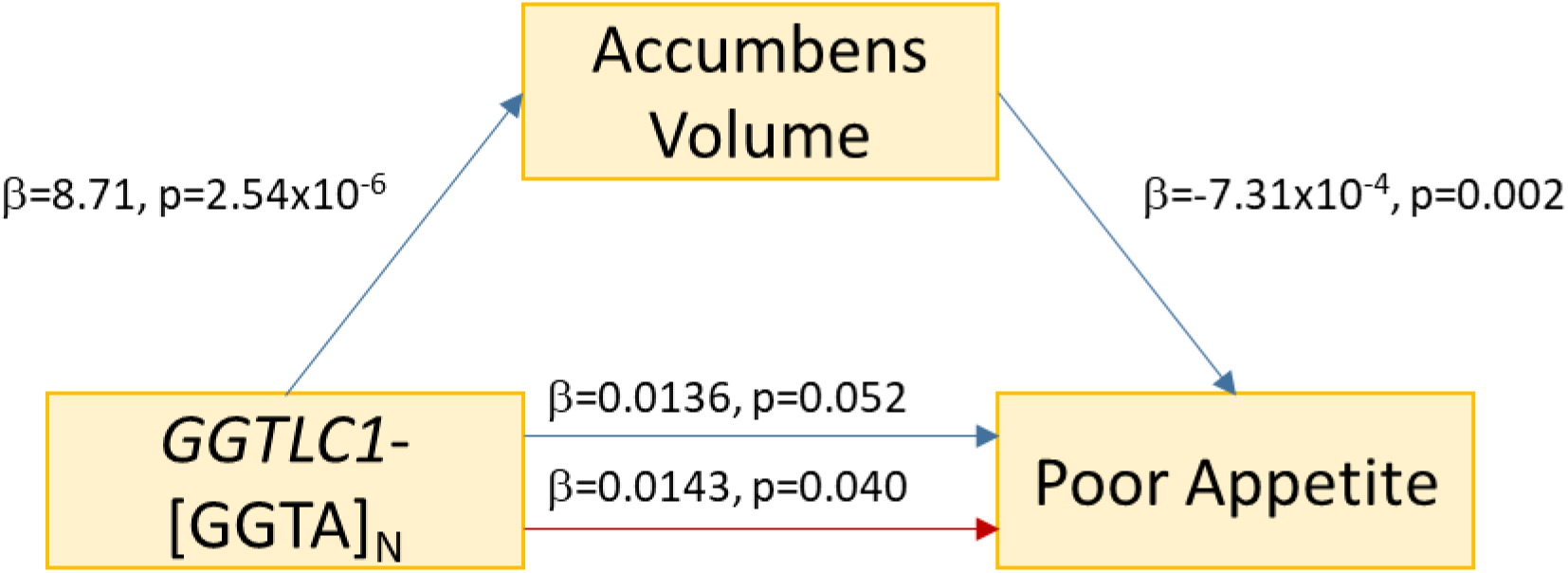
Significant independent effects of *GGTLC1*-[GGTA]_N_ on depression symptoms independent of accumbens volume. Red text denotes the direct effect (ADE) after controlling for the relationship between accumbens volume and poor appetite (UKB Field ID 20511). Refer to Table S9 for additional details for each mediation analysis.

**Figure S10.**
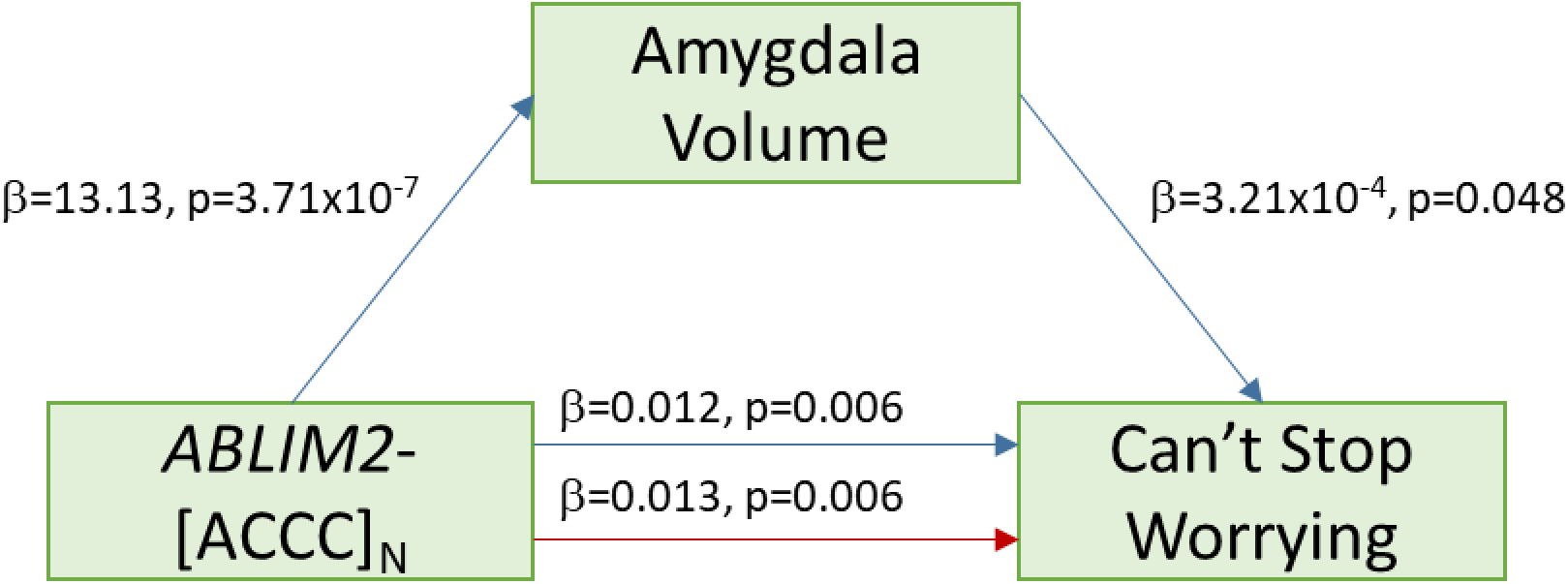
Significant independent effects of *ABLIM2*-[ACCC]_N_ on anxiety symptoms independent of amygdala volume. Red text denotes the direct effect (ADE) after controlling for the relationship between amygdala volume and inability to stop or control worrying (UKB Field ID 20509). Refer to Table S9 for additional details for each mediation analysis.

**Figure S11.**
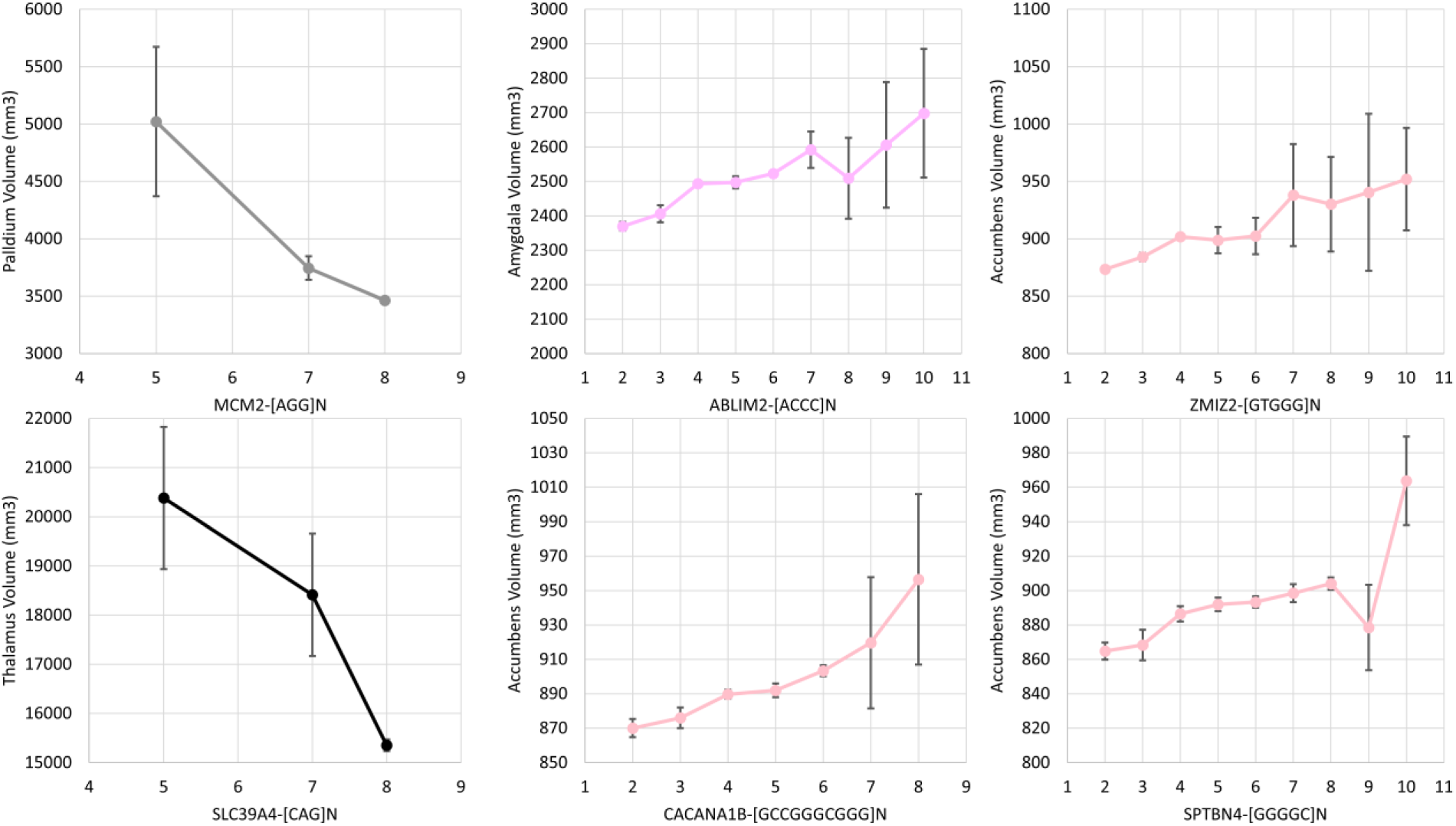
Effects of locus-level burden at the indicated TRs (x-axes) on the labelled subcortical volume (y-axes): amygdala in plum, pallidum in grey, thalamus in black, and accumbens in pink. Error bars per figure reflect the standard error per point estimate.

**Figure S12.**
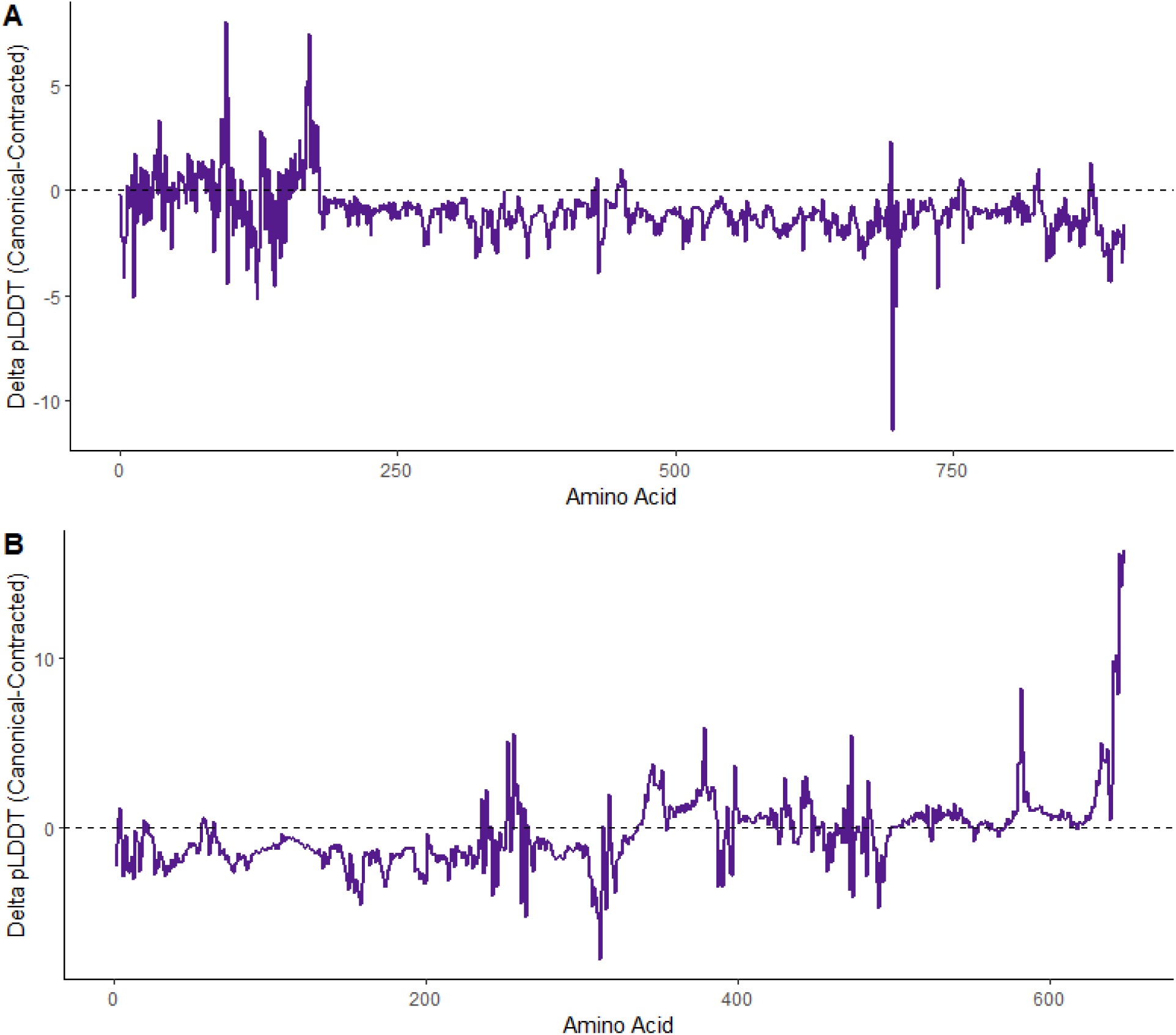
Change in protein structural confidence, measured as ΔpLDDT, between canonical and contracted forms of *MCM2*-[AGG]_N_ (panel A) and *SLC39A4*-[CAG]_N_ (panel B).

### TABLES

**Table S1.** Summary statistics for TRs associated with seven subcortical volumes.

**Table S2.** Fine-mapping summary including number of variants per credible set ("N CS"), posterior inclusion probability (PIP), and whether the TR in that region had the highest PIP.

**Table S3.** Cross-population replication of each TR-subcortical volume association from Table S1. Significant (P<0.05) replications are highlighted in orange. In blue, we highlight nominally significant associations between subcortical volumes and TRs detected in EURs.

**Table S4.** Up, down, and differentially expressed transcriptomic profiles across 54 GTEx v8 tissues, 11 brain developmental stages from BrainSpan, and 29 brain ages from BrainSpan. DEG = differentially expressed gene, adjP = Benjamini-Hochberg adjusted P-value.

**Table S5.** Splicing effects of the *SUPT5H*-[CCA]_N_ intronic tandem repeat in GTEx v8 tissues. Statistical models estimated splicing quantity using TR locus level burden included sex, library preparation protocol, sequencing platform, the first five within-ancestry principal components, and probabilistic estimation of expression residuals.

**Table S6.** Splicing effects of the *SH2B1*-[GCC]_N_ 5’ untranslated region tandem repeat in GTEx v8 tissues. Statistical models estimated splicing quantity using TR locus level burden included sex, library preparation protocol, sequencing platform, the first five within-ancestry principal components, and probabilistic estimation of expression residuals.

**Table S7**. Effects of significantly fine-mapped TRs (Table S2) on relevant subcortical volumes. Two-sided Z-tests were used to compare mean subcortical volumes across TR repeat burden. The magnitude of these differences were captured using Cohen’s *d* estimates.

**Table S8.** Summary of selection coefficients (s) for di-, tri-, and tetra-nucleotide repeats associated with subcortical volumes in the UK Biobank.

**Table S9.** Mediation analysis approach (graphic) and results. The TR effect (X) was derived from our primary association study of TRs versus each brain volume; (Total Effect) reflects the relationship between each TR and the chosen psychiatric symptoms; (ACME) is the effect of brain volume on the psychiatric symptom, and (ADE) is the effect of the TR on each psychiatric symptom independent of the relationship between brain volume and psychiatric symptom. Note that if ACME is not significant, we only report Total Effect of TR on psychiatric symptom. Mediation diagrams are shown in Figures S5-S10.

**Table S10.** Enrichment of pleiotropic signal for each subcortical volume associated gene. Highlighted cells are significant (P<0.05). Gene names with an asterisk (*) denote enrichment performed using nominally significant pleiotropic findings (P<0.05); Gene names without an asterisk denote enrichment performed with pleiotopic findings surviving multiple testing correction (P<2.60x10^-4^, based on 4,756 phenotypes).

